# The association between prenatal and early-life antibiotic use and obesity at 4-5 years in the Born in Bradford birth cohort

**DOI:** 10.1101/2025.04.02.25325113

**Authors:** Lila Goodwin, Ellena Badrick, Gillian Santorelli, Sergio Cunha, Emily Petherick, Sam Oddie, Neil Pearce, John Wright, Lucy Pembrey

## Abstract

**Introduction:** The prevalence of childhood obesity has quadrupled globally over the last 30 years highlighting the urgency for robust research on modifiable risk factors. This study investigates the association between prenatal and early-life (0-24 months) antibiotic use and obesity at 4-5 years within the Born in Bradford (BiB) cohort, and whether this differs by ethnicity or by sex.

**Methods:** BiB is a large, multi-ethnic birth cohort of 13,858 children in Bradford, UK. Data for children and their mothers were obtained from a baseline questionnaire, linked electronic health records (including maternity records and general practitioner (GP) prescriptions), and anthropometric measurements at school. Logistic regression models were employed to estimate the associations between antibiotic exposure in pregnancy and early-life and having obesity at age 4-5 as the outcome, adjusting for several confounders.

**Results:** In this population, nearly a tenth of children had obesity, just over a third were exposed to antibiotics in pregnancy, and nearly three quarters were exposed in the first two years of life. The adjusted analysis revealed no clear evidence of an association between prenatal antibiotic use and having obesity at 4-5 years (aOR 1.09; 95% CI 0.94, 1.27; p= 0.248). However, the associations were stronger in males (aOR 1.24; 95% CI 1.02, 1.52; p= 0.037), and for second trimester exposure (aOR 1.17; 0.97, 1.41). There was strong evidence of an association between early-life antibiotic use and having childhood obesity (aOR 1.36; 95% CI 1.15, 1.62; p=0.0001), with stronger associations in females (aOR 1.58; 95% CI 1.23, 2.02; p=0.0002) and with increasing prescriptions (aOR 1.058; 95% CI 1.029, 1.088; p_trend_=0.0001). No effect modification by ethnicity was observed.

**Conclusion:** These findings support those found in previous experimental animal and epidemiological studies, highlighting the importance of early-life antibiotic use as a potential modifiable risk factor for the development of obesity.

## Introduction

The proportion of children with obesity has been on the rise globally, quadrupling (from 2% to 8%) in the last 30 years^1^. In 2022/2023 in the UK, 9.2% of 4–5-year-olds and 22.7% of 10–11-year-olds were living with obesity, higher than the global average. Obesity in childhood often persists into adulthood, doubling the risk of premature death^2^. Research on the modifiable risk factors for childhood obesity is important to design interventions that can reduce related morbidity and mortality.

The growth-promoting effects of antibiotics were first realised in the 1940s, leading to their low-dose use in livestock farming^3,4^. This was later confirmed by several experimental studies in animal models providing causal evidence for these observed effects^4,5^. Earlier administration (in infancy) has been linked to more extreme weight gain both in agriculture^6^ and in experimental studies in mice, with effects persisting into adulthood^5^.

Antibiotic use is associated with a reduction in gut microbiota diversity, favouring species that extract more energy from food, thus contributing to obesity^7–9^. Other mechanisms may modify or confound the antibiotic-obesity association. For example, the illness motivating the antibiotic prescriptions may result in parental behavioural changes that promote obesity, such as staying inside and not exercising^10^.

Some previous studies found that prenatal antibiotic exposure was associated with an increased risk of having obesity. However, there is no definitive conclusion based on the current evidence, with several studies finding null associations and inconsistency in the magnitude of effect^11–15^. The largest effect size was reported by a prospective cohort study in Greece, where prenatal self-reported antibiotic use was associated with twice the odds of living with obesity at 6 years (RR 2.09; 95% CI 1.58, 2.76)^13^. Only two previous studies investigated effect modification by sex^16,17^, but neither found evidence of this. Few studies have investigated differences by ethnicity, either because of predominantly White populations or small sample sizes.

There is strong evidence of an association between early-life (0-4 years) antibiotic use and childhood obesity, with several meta-analyses yielding positive associations^18–20^. The effect sizes for individual studies ranged from 1.04-6.15^11,18–22^. The largest effect size was reported by a prospective cohort study in California, USA (aOR 6.15; 95% CI 1.03, 36.70), although wide CIs indicate some imprecision likely due to a small sample size (97 children)^22^. There is some evidence of a dose-response effect, with increasing number of prescriptions associated with greater odds of having childhood obesity^18^. Only one study reported the findings by ethnicity and found a weak overall association between antibiotic use and obesity (aOR 1.1; 95% CI 0.6, 1.8), but strong evidence of an association in Hispanic children (aOR 3.3; 95% CI 1.7, 6.4)^23^. Of the studies that investigated this association by sex, most found stronger associations in boys, suggesting potential sexual dimorphism, though the evidence for this is inconsistent^24–26^.

While existing research has laid a foundation for understanding the association between antibiotic use and obesity, gaps remain, especially in understanding effect modification by ethnicity and sex. Several previous studies experienced problems associated with selection bias^13^, small sample size^22^ and small numbers of children with obesity^21^. This study addresses these limitations by investigating the association within the BiB cohort, a large multi-ethnic birth cohort. Children of Pakistani ethnicity have one of the highest prevalences of severe obesity in the UK^27^, making research in this group a priority. To our knowledge, this study is the first to explore this association in children of Pakistani heritage.

## Methods

### Cohort information

BiB is a multi-ethnic prospective, longitudinal birth cohort established in 2007 in Bradford, West Yorkshire; nearly 50% of babies born there are of Asian/Asian British ethnicity (90% of which are of Pakistani heritage). Bradford is one of the most deprived areas in England.

Between September 2007 and December 2011, 12,453 women (13,776 pregnancies) were recruited from Bradford Royal Infirmary at 26-28 weeks’ gestation, resulting in 13,858 births, which are representative of all births in Bradford^28^. Women who agreed to participate signed a consent form.

### Sources of data

Data on ethnicity, socio-economic factors and maternal behaviours during pregnancy were self-reported in a baseline questionnaire administered at recruitment. The population analysed was restricted to Asian/Asian British Pakistani and White British individuals. Ethnicity was defined as ethnic group (e.g. Asian/Asian British, White British) and cultural background (e.g. Pakistani, Bangladeshi, Indian). Socio-economic position (SEP) was defined using a BiB specific latent class analysis of 19 component variables^29^. Early pregnancy/pre-pregnancy body mass index (BMI) was derived from height measured at pregnancy booking and weight measured at recruitment to BiB. Information on antibiotic prescriptions were extracted from linked electronic health records (supplementary material 1). Birth data was obtained from the hospital maternity records. Child BMI at 4-5 years was sourced from the National Child Measurement Programme (NCMP) in which school nurses measured the height and weight of children in reception (aged 4-5) using calibrated SECA scales and consistent operating procedures.

### Exposure definitions

Prenatal antibiotic exposure was categorised as a binary variable, where children were considered as exposed if their mothers received any antibiotic prescriptions during pregnancy (see supplementary material 1 for prescription code list). This was further divided into 3 binary non-mutually exclusive variables corresponding to any prescriptions in each trimester: first trimester (<93 days of gestational age); second trimester (93–184 days); third trimester (185 days of gestational age to 7 days before the child’s birth). For the BiB cohort and in the UK generally, gestational age is estimated from the date of delivery and the 10-week ultrasound scan. Early-life antibiotic exposure was defined as both a binary variable of any antibiotic prescriptions between 0-24 months of age, and as a continuous variable of the total number of prescriptions during this period.

### Outcome definition

The outcome, having obesity at 4-5 years, was analysed as a binary variable (children with obesity/children not living with obesity), based on BMI z-scores. BMI was calculated from weight and height measurements taken by nurses using the following formula:

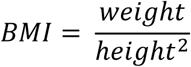

Standardised BMI z-scores were calculated for each child’s BMI using the LMS method (LMS growth, Health for all children) based on the UK90 reference data^30,31^, which is recommended for children aged 4 and over and is used by the NCMP^27^. Children were defined as living with obesity if they had a BMI z-score above the 95^th^ percentile^32^. For the baseline characteristics and sensitivity analysis, living with overweight was defined as a BMI z-score above the 85^th^ percentile, up to the 95^th^ percentile^32^.

### Covariate definitions

For mother’s BMI, an adjustment was applied to measurements taken at >13 weeks’ gestation (2,580 mothers), as women begin to experience gestational weight gain at that time^33^. This adjustment was based on an average weight gain of 12.48kg between weeks 14-40 of gestation^33^ as found in previous studies, assuming a linear relationship between week of gestation and weight gain (0.48kg per week). The same adjustment was applied to all women due to a lack of research on trajectories by ethnicity:

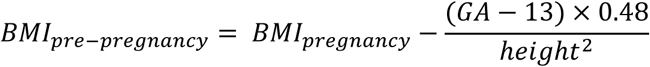

This adjustment resulted in a reduction in mean BMI for the 2,580 women who had their BMI measured past 13 weeks from 26.35 (SD 5.61) to 25.39 (SD 5.56), reflecting a more accurate pre-pregnancy BMI. Pre-pregnancy BMI was categorised as a binary variable (mothers with pre-pregnancy obesity/mothers without pre-pregnancy obesity). Mothers were defined as living with obesity if they had a pre-pregnancy BMI of ≥30 kg/m2^34^ for White British women, and BMI of ≥27.5kg/m2 for Pakistani women, based on updated cut-offs for Asian (including Pakistani) individuals^35^.

### Statistical analysis

All analyses were performed using Stata SE 18.0.

The study population’s characteristics were summarised, stratified by obesity status and ethnicity. Baseline characteristics of those missing obesity and antibiotic data were compared to those without missing data to assess selection bias. Sex (assigned at birth and classified as male or female) and ethnicity were included *a priori* in all models. All explanatory variables in table 1 were entered as potential confounders and were included in the regression models if they met three criteria (1) they were a risk factor for obesity (assessed through univariate analysis), (2) they were independently associated with prenatal and/or early-life antibiotic exposure (assessed through cross-tabulations) and (3) they were not on the casual pathway (assessed through Directed Acyclic Graphs (Figure 1))^36,37^.

**Figure 1:**
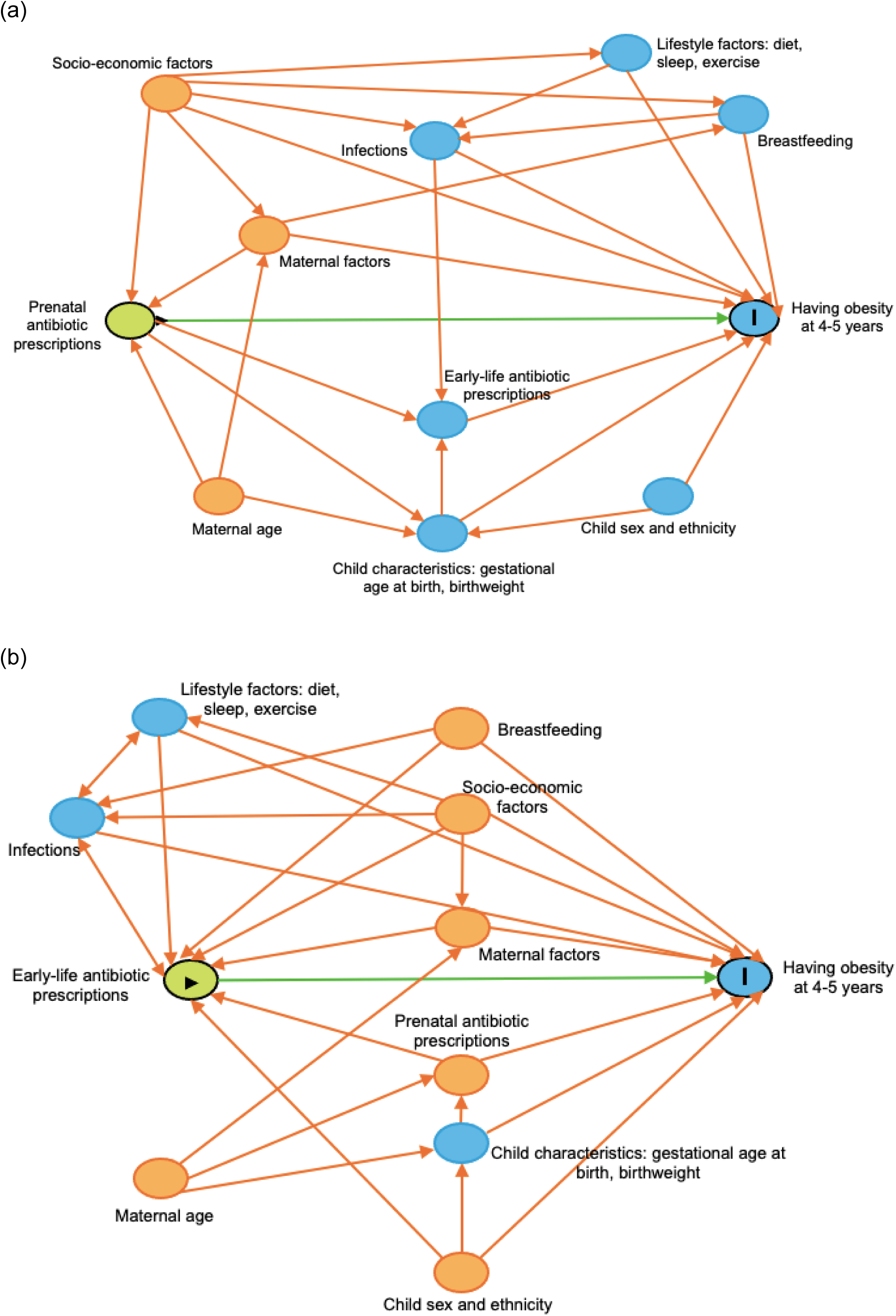
Directed acyclic graphs for the association between. (a) prenatal and (b) early-life antibiotic use and childhood obesity, based on previous research. Designed in Dagitty^37^. Green with triangle= exposure, blue with I= outcome, orange= potential confounders, blue= other variables. Note maternal factors includes smoking during pregnancy, gestational diabetes, pre-pregnancy obesity and parity.

**Table 1:**
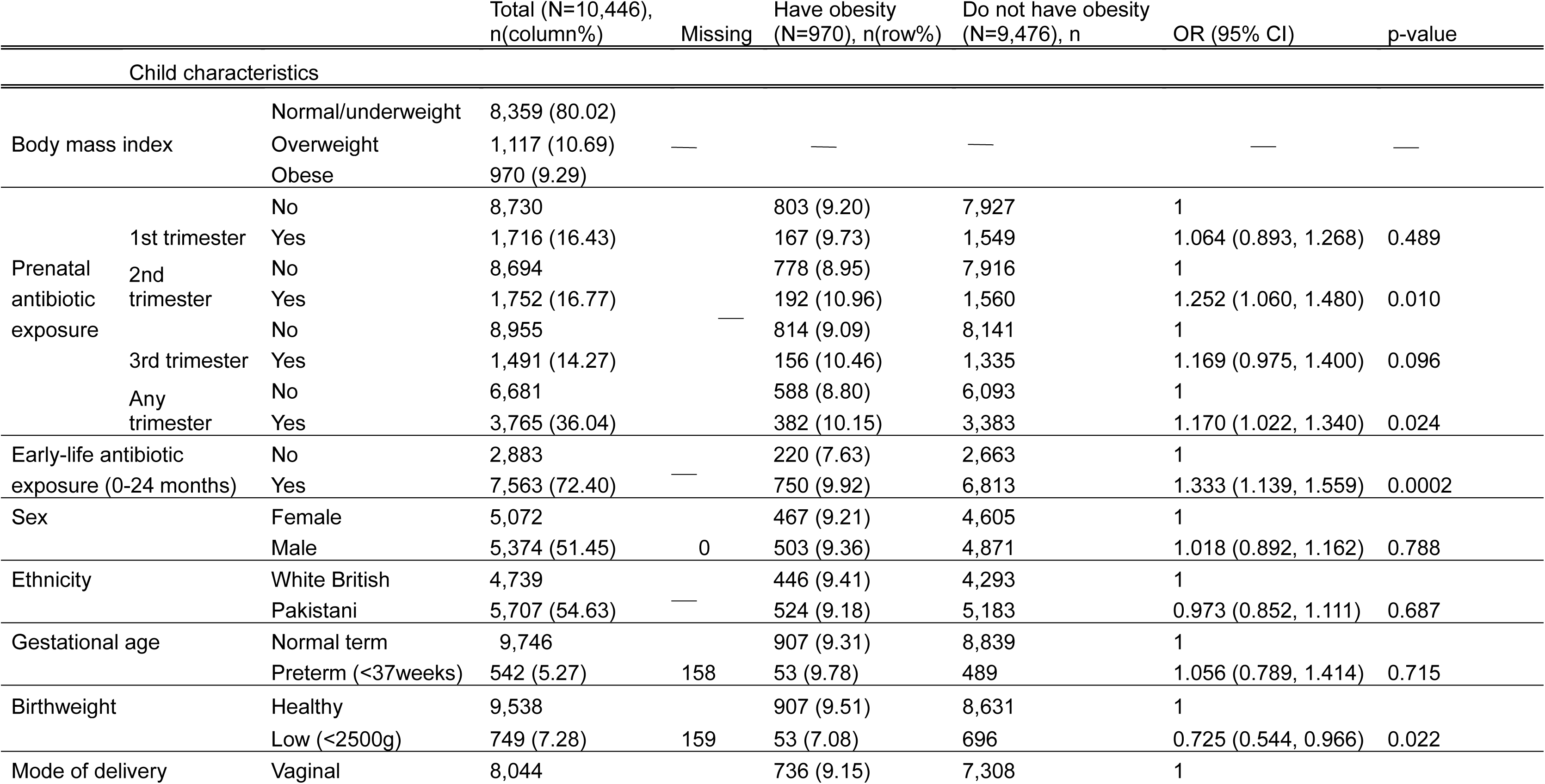
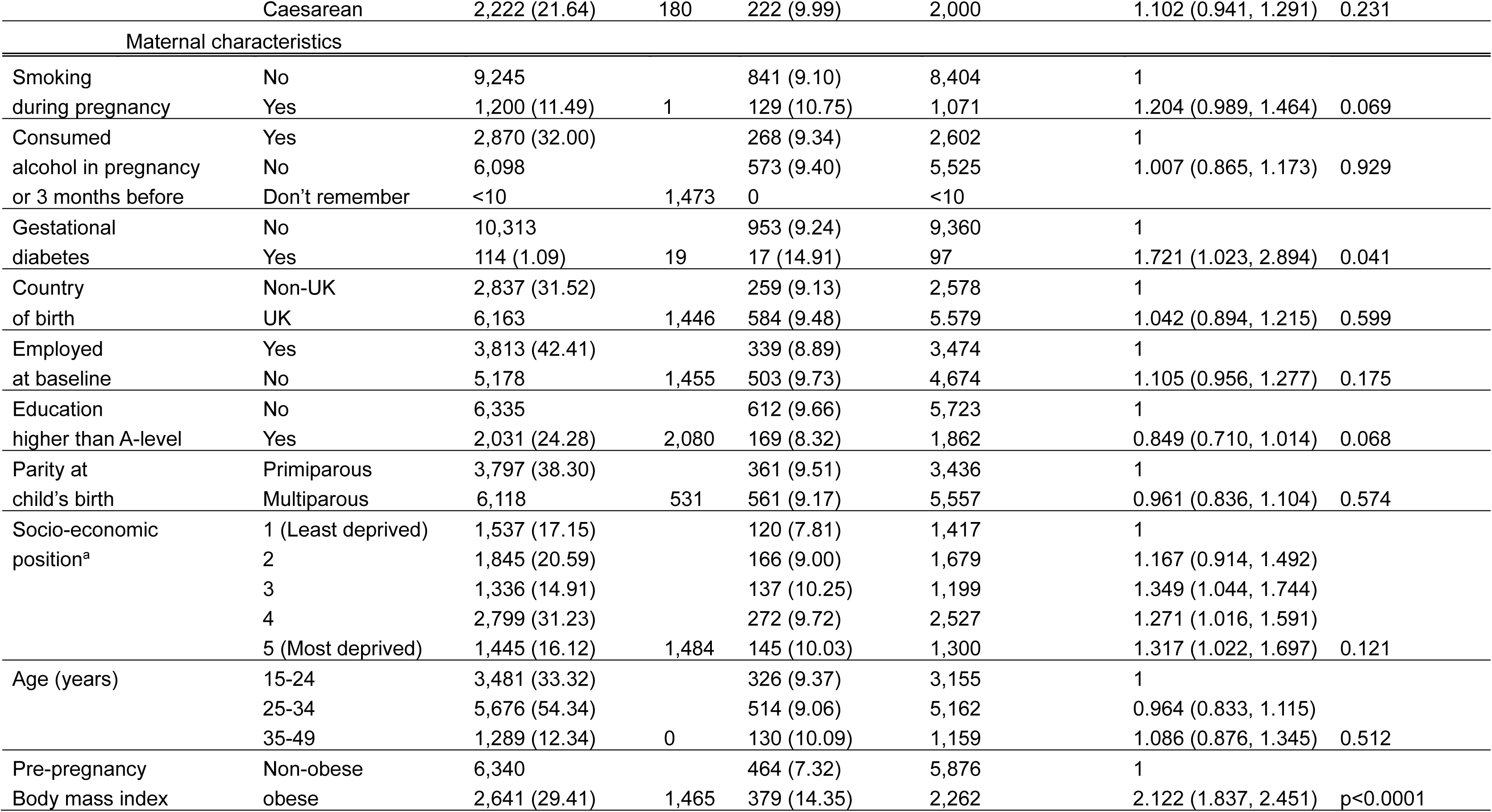
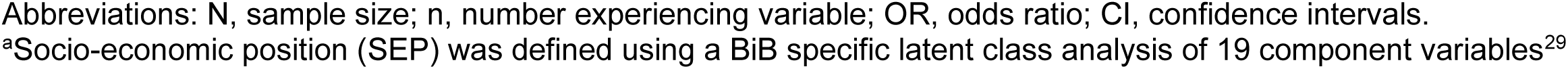
Baseline characteristics of the study population (N=10,446) with stratification by obesity at 4-5 years; ORs and 95% CIs quantifying the association between each variable and obesity are from univariate logistic regression models and p-values from Likelihood Ratio Tests.

Confounders were investigated separately for prenatal and early-life exposure. Prior to model building, a crude analysis with each exposure, outcome, and *a priori* confounders was conducted to obtain crude odds ratios (ORs), 95% CIs and Likelihood ratio test (LRT) p-values.

### Multivariate analysis

A backward modelling approach was used. All potential confounders that met the criteria were added to their respective logistic regression models as per model specifications (Figure 2). The standard error of each full model was compared to that of the crude model. A large increase in standard error of the OR (>10%) suggested potential collinearity between covariates, making the model unstable. If this occurred the pseudo-R^2^ was calculated for models with and without the potentially collinear variable to determine whether it should be retained based on its collinearity and confounding effect^36^.

**Figure 2:**
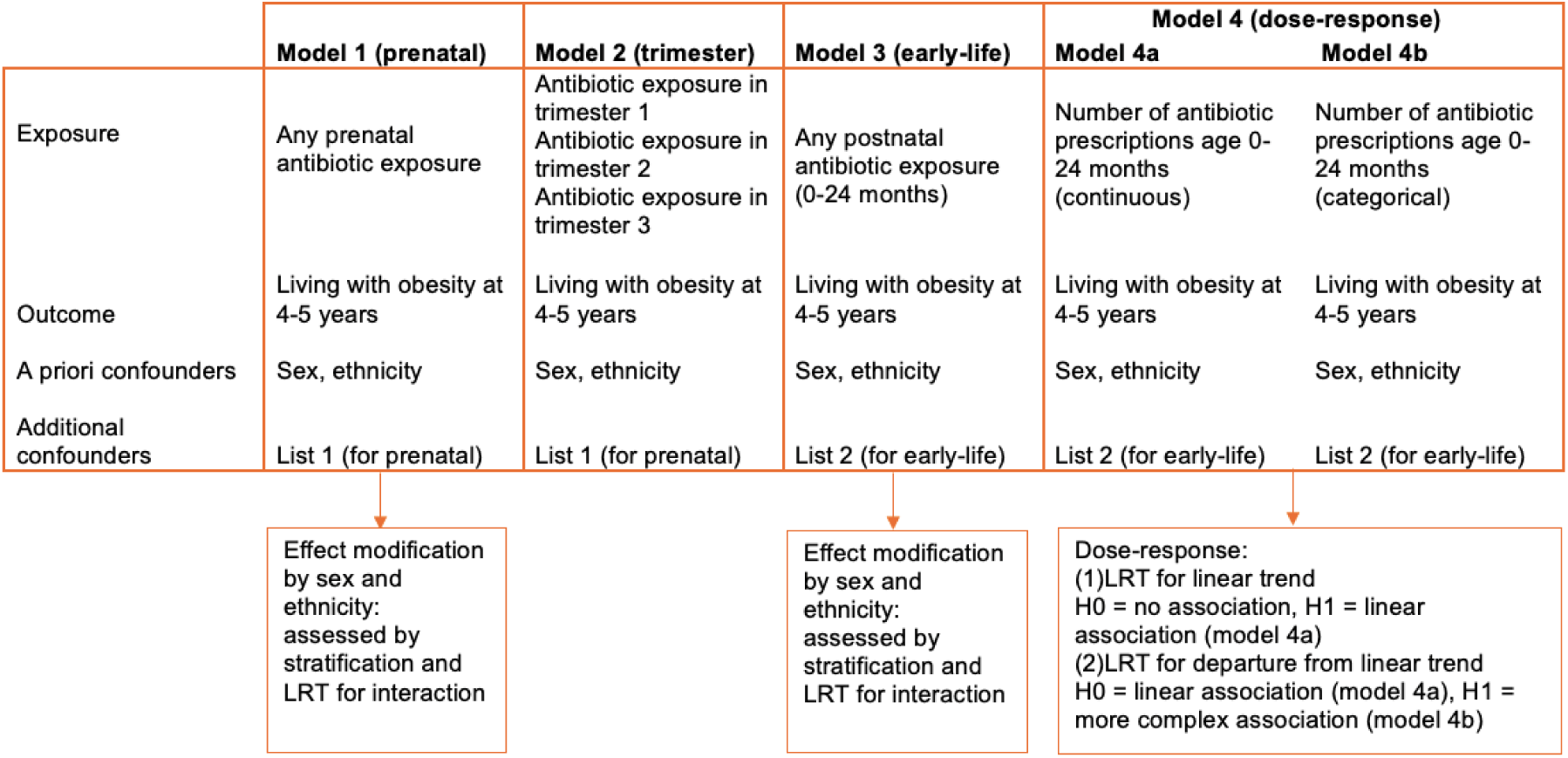
Model specifications and tests to be run on each model. Refer to section 3.4 for confounder list 1 and 2. Abbreviations: LRT, likelihood ratio test; H0, null hypothesis; H1, alternate hypothesis.

### Sensitivity analysis

All siblings were included in the main analysis to maximize power and generalizability, as there is evidence that risk of obesity varies with parity^38^. A sensitivity analysis with only first siblings (N=9,371) was conducted to confirm results with truly independent observations. Some studies have shown that obesity estimated by body fat percentage was similar to overweight/obesity estimated by BMI^39^. Overweight is also the precursor to obesity and is associated with adverse health outcomes^1^. Therefore, a second sensitivity analysis included children with overweight (z-scores≥85th percentile) in the outcome. Previous studies have shown that breastfeeding could confound the association between antibiotic use and having obesity^21,26,40^. Breastfeeding data were not available for all children so a subgroup analysis (n= 8,991) was conducted to investigate the effect of any breastfeeding in this population (supplementary material 2). This analysis was limited to early-life antibiotic exposure as breastfeeding is likely on the causal pathway between prenatal antibiotic use and obesity (Figure 1).

### Ethical approval

The original BiB study received ethical approval from the Bradford Research Ethics Committee (Ref 07/H1302/112). The NIHR antibiotics study has Health Research Authority (HRA) and Health and Care Research Wales (HCRW) approval (ref: 238908). Ethical approval for this study was granted by the London School of Hygiene and Tropical Medicine ethics committee (Ref 30315).

## Results

### Selection of study population

The study population included all singleton alive births in the BiB cohort that had complete BMI, antibiotic exposure, and ethnicity data and were of Pakistani or White British ethnicity (N=10,446) (Figure 3).

**Figure 3:**
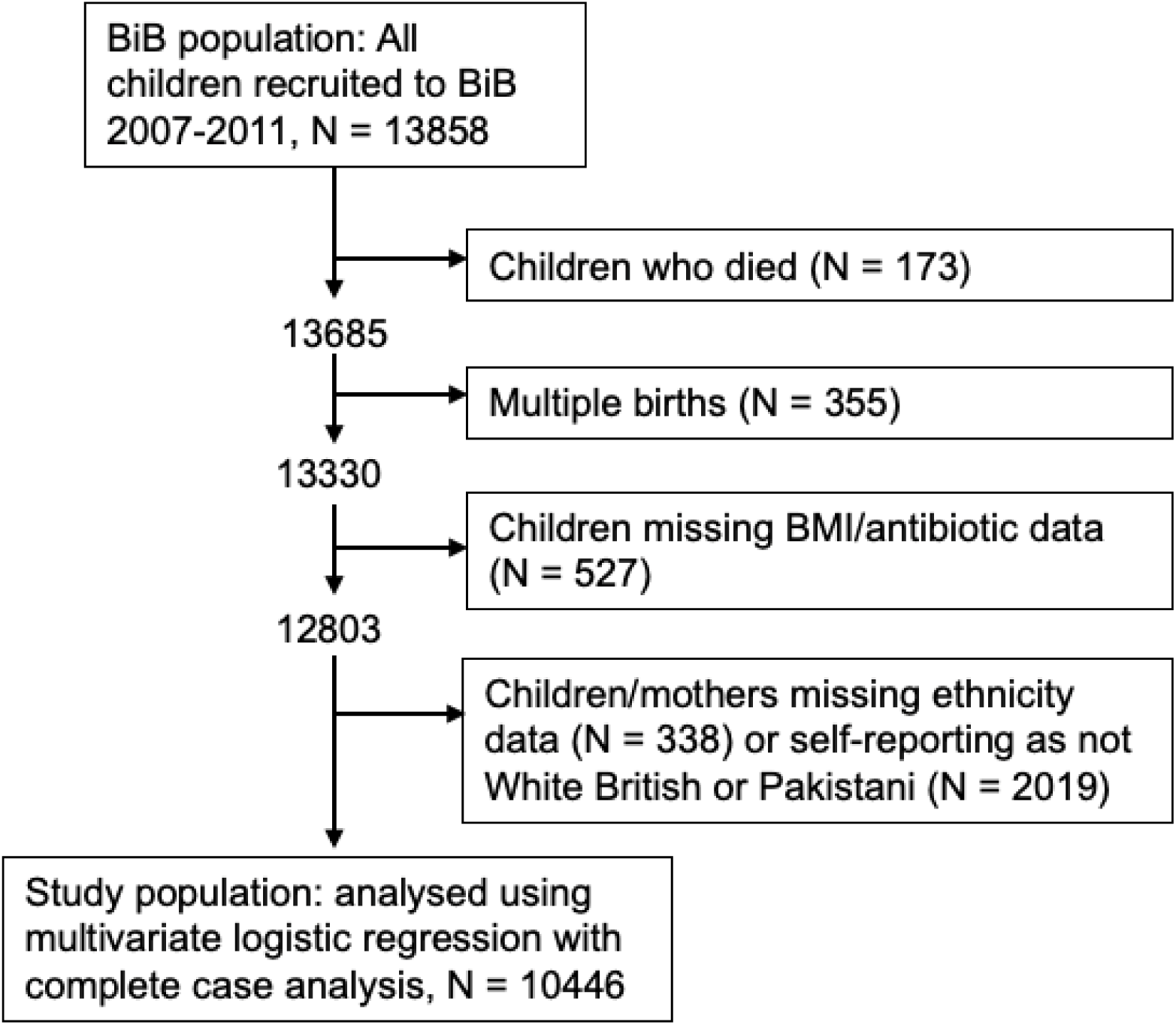
The selection process of the study population.

### Baseline characteristics

Nearly half of the population were in the most socioeconomically disadvantaged groups. In this study, 72% of children were exposed to antibiotics in their first 2 years of life while 36% were exposed prenatally (Table 1). Nearly a tenth of children were classified as having obesity (n=970) with a further 1,117 (11%) classified as having overweight. Having obesity was more common in those prenatally exposed to antibiotics (10%) and those exposed in early-life (10%), than those who were unexposed (9% and 8%, respectively) (Table 1).

Obesity was more prevalent in children of mothers who smoked during pregnancy (11%), had gestational diabetes (14%), had pre-pregnancy obesity (14%) or were from more deprived SEPs, compared to the overall study population (9%) (Table 1). The study population is broadly representative of the larger BiB cohort (supplementary material 3).

Over half of children were of Pakistani ethnicity. A fifth of White British women smoked during pregnancy and 68% consumed alcohol during pregnancy or three months prior, compared to only 3% of Pakistani women who smoked and less than 1% who drank alcohol. Gestational diabetes was twice as prevalent in Pakistani women compared to White British. Pre-pregnancy obesity was also more common in Pakistani mothers (32%) compared to White British (26%). For further baseline characteristics stratified by ethnicity see supplementary material 3.

### Univariate analysis

Confounding variables were determined to be: For prenatal antibiotic exposure: gestational diabetes, SEP, smoking during pregnancy, maternal pre-pregnancy obesity. For early-life antibiotic exposure: low birthweight, SEP, smoking during pregnancy, maternal pre-pregnancy obesity, prenatal antibiotic use.

### Multivariate analysis

#### Prenatal antibiotic exposure

In the crude logistic regression model, prenatal antibiotic exposure was associated with 1.17 times the odds of obesity (95% CI 1.02, 1.34) (Table 2). After adjusting for all additional confounders there was no evidence of an overall association (aOR 1.09; 95% CI 0.94, 1.27; p=0.248).

**Table 2:** Association between prenatal antibiotic exposure and obesity at 4-5 years (N=10,446), stratified by ethnicity (Pakistani and White British) and sex.

|  | Crude OR* (95% CI), p-value | aOR** (95% CI), p-value | p-value<sup>i</sup> |
| --- | --- | --- | --- |
| All | 1.171 (1.022, 1.341), 0.023 | 1.091 (0.941, 1.265), 0.248 |  |
| Stratified by ethnicity |  |  |  |
| White British | 1.218 (0.997, 1.488), 0.054 | 1.081 (0.871, 1.342), 0.482 | 0.892 |
| Pakistani | 1.132 (0.942, 1.362), 0.187 | 1.084 (0.885, 1.328), 0.435 |  |
| Stratified by sex |  |  |  |
| Female | 1.060 (0.871, 1.291), 0.561 | 0.940 (0.758, 1.166), 0.572 | 0.105 |
| Male | 1.283 (1.064, 1.547), 0.010 | 1.244 (1.015, 1.524), 0.037 |  |

ORs, aORs and 95% CIs are from logistic regression models; p-values are from Likelihood Ratio Tests (testing null hypothesis of no association).
\*Adjusted for *a priori* confounders, sex and ethnicity
\*\*Additionally adjusted for diabetes, maternal pre-pregnancy obesity, socio-economic position, smoking during pregnancy.
<sup>i</sup>from Likelihood Ratio Tests for interaction with the aORs.
Abbreviations: N, sample size; OR, odds ratio; aOR, adjusted odds ratio; CI, confidence interval.

When analysed by ethnicity, there was no evidence of an association in either White British (aOR 1.08, 95% 0.87, 1.34; p=0.482) or Pakistani children (aOR 1.08, 95% CI 0.89, 1.33; p=0.435) (Table 2), and no evidence for effect modification (p=0.892). However, stratification by sex showed some evidence of an association between prenatal antibiotic use and obesity in male children (aOR 1.24, 95% CI 1.02, 1.52; p=0.037), while no association was seen in female children (aOR 0.94, 95% CI 0.76, 1.17; p=0.572) (Table 2). A formal LRT showed some evidence of effect modification by sex (p=0.105).

There was evidence of a crude association between antibiotic exposure in the second trimester and obesity at 4-5 years, OR 1.23 (95% CI 1.03, 1.46), but not the first (OR 1.02; 95% CI 0.85, 1.22) or third trimester (OR 1.12; 95% CI 0.93, 1.35) (Table 3). After adjustment, the ORs for all trimesters decreased, and there was weak evidence (p=0.099) for an association between second trimester antibiotic exposure and obesity (aOR 1.17; 95% CI 0.97, 1.41) (Table 3).

**Table 3:**
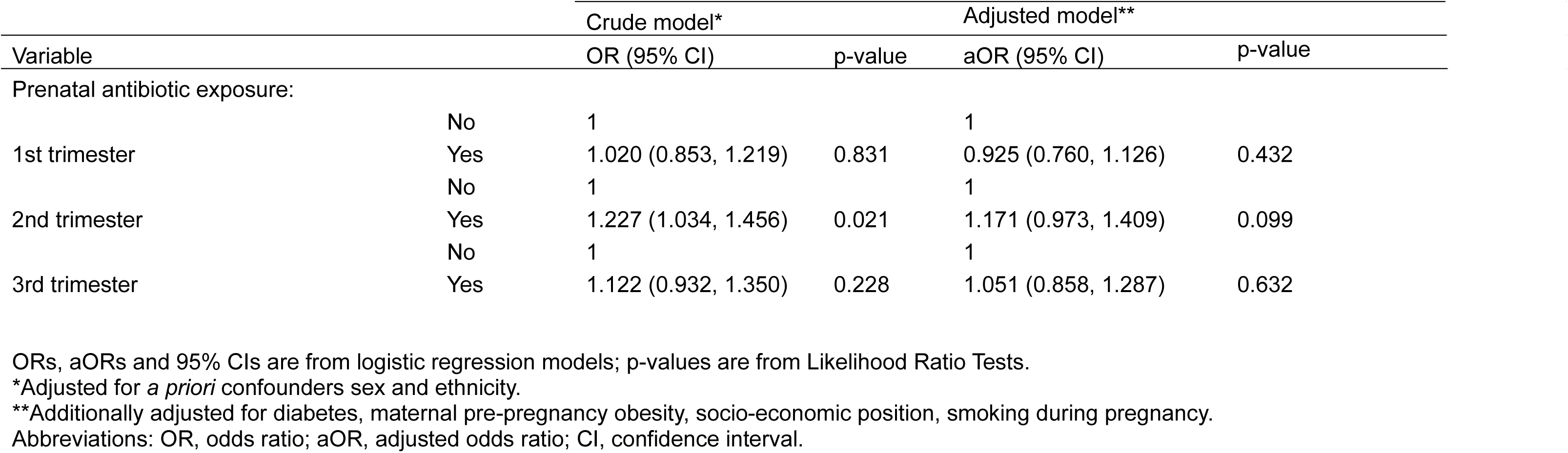
Association between prenatal antibiotic exposure in each trimester and obesity at 4-5 years among 10,446 children.

|  |  | Crude model* |  | Adjusted model** |  |
| --- | --- | --- | --- | --- | --- |
| Variable |  | OR (95% CI) | p-value | aOR (95% CI) | p-value |
| Prenatal antibiotic exposure: |  |  |  |  |  |
| 1st trimester | No | 1 |  | 1 |  |
|  | Yes | 1.020 (0.853, 1.219) | 0.831 | 0.925 (0.760, 1.126) | 0.432 |
| 2nd trimester | No | 1 |  | 1 |  |
|  | Yes | 1.227 (1.034, 1.456) | 0.021 | 1.171 (0.973, 1.409) | 0.099 |
| 3rd trimester | No | 1 |  | 1 |  |
|  | Yes | 1.122 (0.932, 1.350) | 0.228 | 1.051 (0.858, 1.287) | 0.632 |

ORs, aORs and 95% CIs are from logistic regression models; p-values are from Likelihood Ratio Tests.
\*Adjusted for *a priori* confounders sex and ethnicity.
\*\*Additionally adjusted for diabetes, maternal pre-pregnancy obesity, socio-economic position, smoking during pregnancy.
Abbreviations: OR, odds ratio; aOR, adjusted odds ratio; CI, confidence interval.

#### Early-life antibiotic exposure

There was strong evidence of a crude (OR 1.34; 95% CI 1.14, 1.57; p<0.0001) and adjusted (aOR 1.36; 95% CI 1.15, 1.62; p=0.0001) (Table 4) association between early-life antibiotic use and obesity.

**Table 4:** Association between early-life antibiotic exposure and obesity at 4-5 years (N=10,446), stratified by ethnicity (Pakistani and White British) and sex.

|  | Crude OR* (95% CI), p-value | aOR** (95% CI), p-value | p-value<sup>i</sup> |
| --- | --- | --- | --- |
| All | 1.339 (1.144, 1.568), <0.0001 | 1.362 (1.145, 1.619), 0.0001 | — |
| Stratified by ethnicity |  |  |  |
| White British | 1.312 (1.054, 1.634), 0.014 | 1.341 (1.059, 1.699), 0.013 | 0.828 |
| Pakistani | 1.368 (1.090, 1.718), 0.006 | 1.404 (1.087, 1.812), 0.008 |  |
| Stratified by sex |  |  |  |
| Female | 1.403 (1.126, 1.749), 0.002 | 1.579 (1.233, 2.021), 0.0002 | 0.146 |
| Male | 1.274 (1.016, 1.596), 0.032 | 1.185 (0.929, 1.512), 0.166 |  |

Crude, aORs and 95% CIs are from logistic regression models, and p-values from Likelihood Ratio Tests (testing null hypothesis of no association).
\* Adjusted for *a priori* confounders sex and ethnicity
\*\*Additionally adjusted for maternal pre-pregnancy obesity, birthweight, socio-economic position, smoking during pregnancy, prenatal antibiotic use.
<sup>i</sup>from Likelihood Ratio Test for interaction with the aORs.
Abbreviations: N, sample size; OR, odds ratio; aOR, adjusted odds ratio; CI, confidence interval.

Stratification of the adjusted model (Model 3) by ethnicity showed a slightly stronger association between early-life antibiotic use and obesity in Pakistani children (aOR 1.40, 95% CI 1.09, 1.81; p=0.008) compared to White British children (aOR 1.34, 95% CI 1.06, 1.70; p=0.013) (Table 4). A formal LRT presented no evidence to suggest that ethnicity modifies this association (p=0.828). Stratification by sex showed that female children prescribed antibiotics between 0-24 months of age had 1.58 times the odds of obesity compared to unexposed female children (95% CI 1.23, 2.02; p=0.0002). In males, the association was weaker (aOR 1.19; 95% CI 0.93, 1.51; p=0.166) (Table 4), although a formal LRT for interaction did not provide good evidence for effect modification by sex (p=0.146).

Children receiving ≥10 antibiotic prescriptions between 0-24 months of age had the highest proportion of obesity (Table 5). There was strong evidence of a dose-response relationship between the number of early-life antibiotic prescriptions and odds of obesity after adjusting for confounders (p-value for linear trend = 0.0001). Each additional prescription was associated with a 5.8% increase in odds of obesity (95% CI 2.9%, 8.8%) (Table 5). When examined as a categorical variable, every number of antibiotic prescriptions (except 8) was associated with increased odds of obesity, although sample size and precision decreased with increasing numbers of prescriptions. There was no evidence to suggest a more complex (non-linear) association (p-value for departures from linear trend = 0.295) (Table 5).

**Table 5:**
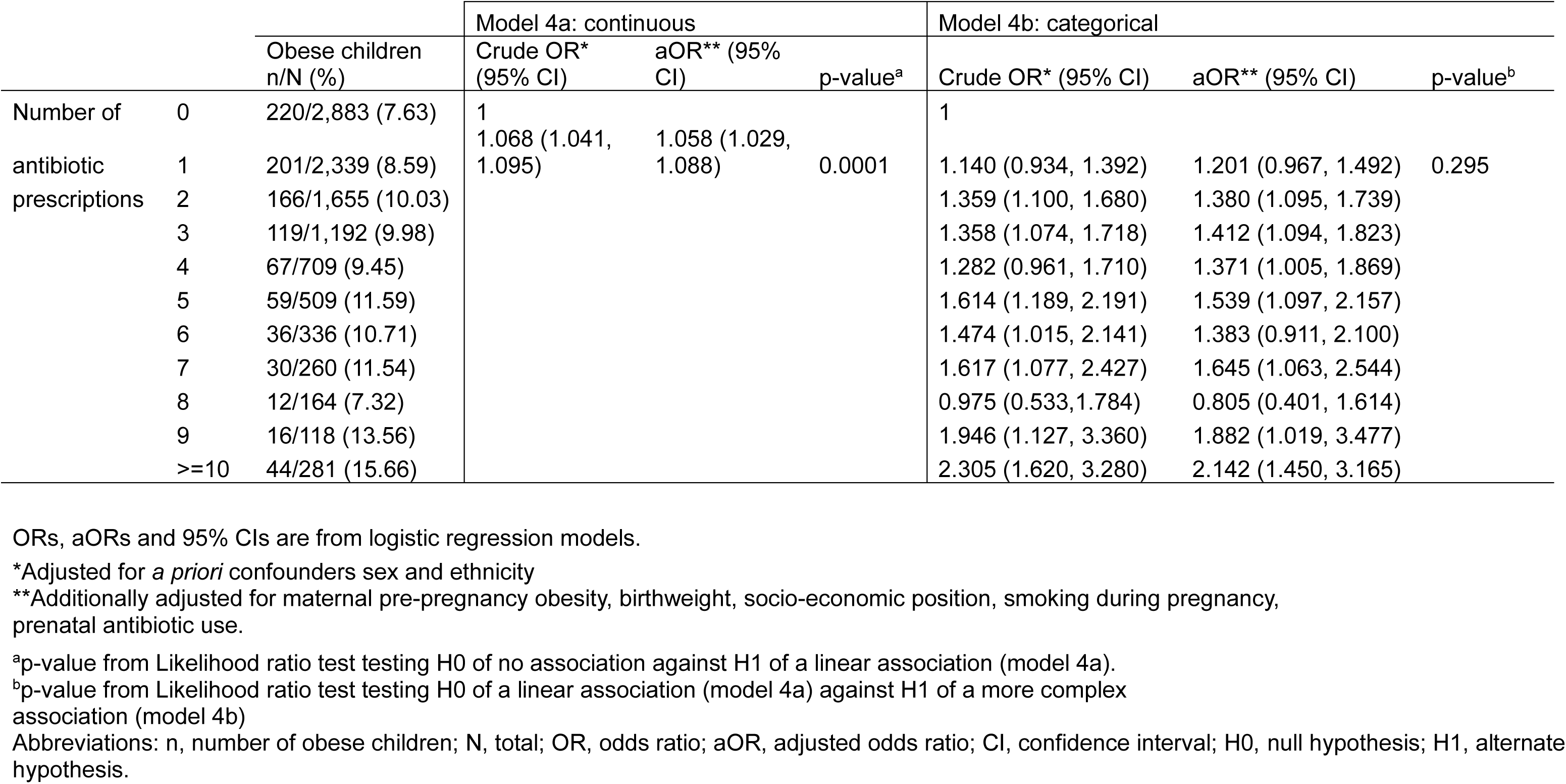
Association between number of antibiotic prescriptions (between 0-24 months of age) and obesity at age 4-5, N=10,446.

### Sensitivity analysis

See supplementary material 5 for the sensitivity analysis result tables. The sensitivity analysis including only first siblings confirmed the main analysis findings for prenatal and early-life exposure.

With obesity/overweight as the outcome there was weak evidence of an association with prenatal antibiotic exposure with narrower 95% CI than the main analysis (aOR 1.11; 95% CI 1.00, 1.24). There was some evidence for effect modification by ethnicity (p=0.02) with evidence of an association in Pakistani children (aOR 1.26; 95% CI 1.08, 1.46; p=0.003) but not White British children (aOR 0.97; 95% CI 0.83, 1.13; p=0.691). The results for effect modification by sex and by trimester findings were the same as the main analysis, with stronger associations found with second trimester exposure (aOR 1.14; 95% CI 1.00, 1.31; p=0.059). The results mirrored the main analysis for early-life antibiotic exposure including strong evidence for an association between early-life antibiotic exposure and having overweight/obesity at 4-5 years (aOR 1.22; 95% CI 1.08, 1.37; p=0.002) and strong evidence for a dose-response relationship (p<0.0001) (the latter is not shown in supplementary material).

In the subgroup analysis (n=8,990) breastfeeding was not associated with early-life antibiotic use so did not fulfil the criteria to be included as a confounder in this population. Although even when included as a confounder the results confirm that of the main analysis with early-life antibiotic use being strongly associated with childhood obesity (aOR 1.36; 95% CI 1.15, 1.62; p<0.001) (supplementary material 2).

## Discussion

### Summary of findings

Overall, there was no clear evidence for an association between prenatal antibiotic exposure and obesity at 4-5 years. Among male children, having at least one antibiotic prescription prenatally was associated with 1.24 times the odds of having obesity (p=0.037), although a formal LRT for interaction suggests this may be due to chance (p=0.105). There was no effect modification by ethnicity. Children receiving at least one antibiotic prescription between 0-24 months of age had 1.36 times the odds of obesity at 4-5 years compared to children who did not receive any prescriptions (p=0.0001). This association was stronger in females, with early-life antibiotic exposure associated with 1.58 times the odds of obesity (p=0.0002).

There was no evidence of effect modification by ethnicity. Early-life prescriptions had a dose-response effect on obesity, with each additional antibiotic prescription being associated with a 5.8% increase in odds of having obesity.

### Interpretation

Obesity prevalence in this cohort is 9.3%, in line with the national average for 2015 of 9.1%^41^. The results support previous studies in the USA, Japan and China^15–17^, finding no evidence of an association between antibiotic use in pregnancy and having obesity in childhood. Three studies that found associations between antibiotic use in pregnancy and obesity measured obesity at age 7 or later^15^ indicating there may be longer term effects of antibiotic use on obesity not captured in our study. We observed the strongest association with antibiotic exposure in the second trimester. This has been found by previous studies, including a large cross-sectional study in New Zealand^11^ and a prospective cohort study in California^14^. The tunica mucosa layer of the gut forms during the second trimester^42^ which could influence gut microbiome vulnerability however this assumes gut colonisation begins in utero despite evidence for the sterile womb hypothesis^43^.

Our study, in line with several systematic reviews and an observational study in the UK^18–20^, found strong associations between early-life antibiotic use and obesity. One previous study that did not find associations used BMI self-reported from parents^18^. Parental self-reported BMI has been shown to underestimate the weight of heavier children and overestimate the weight of lighter children^44^, which could underestimate the true association.

A causal association between early-life antibiotic use and obesity at age 4-5 is plausible as we have adjusted for the important confounders in the DAG (Figure 1). The association between antibiotic use and childhood obesity is likely mediated via the gut microbiome.

Experimental animal studies have shown that a faecal microbiota transplant (FMT) from an adult female human with obesity^45^ to germ-free mice induced an obese phenotype in the mice. Observational human studies have shown that antibiotic use reduces gut microbiome diversity and favours species that are more energy efficient, such as Firmicutes^7–9^. A 20% increase in Firmicutes could result in 150kcal more energy extracted per day^9^ contributing to increased fat deposition. Some studies suggest gut microbiome establishment begins in utero^46^, however this study suggests it is most susceptible to the effects of antibiotics in the first two years of life, likely due to direct exposure rather than prenatal exposure though the mother. Early-life antibiotic use could also be a proxy for the infection which they are treating. Previous studies found infection is bi-directionally associated with obesity, mediated by inflammation^47^, which could partially explain this observed association.

In this study the association between prenatal antibiotic exposure and having obesity was stronger in males. Previous studies in Japan and China, that measured obesity at 1-4 years, found higher effect sizes for females^16,17^. The observed differences may be due to variation between countries/populations or due to chance.

In the association between early-life antibiotic use and obesity, stratification by sex showed a stronger association in females. Biologically, females may be more vulnerable to antibiotic-induced gut microbiota changes, as studies have observed a higher Firmicutes to Bacteroidetes ratio in females from birth^48^ which is associated with obesity and antibiotic use^9^. Most previous studies that investigated effect modification by sex, found stronger associations in boys, although none adjusted for prenatal antibiotic exposure, which could partially explain this difference.

To our knowledge, this study was the first to investigate this association in children of Pakistani heritage and it appears to not modify the association despite children aged 4-5 years of Pakistani heritage being at increased risk of severe obesity in the UK^27,41^.

### Strengths

This study has several strengths. Its large sample size provides sufficient power to detect effect modification by sex and ethnicity; flexible recruitment strategies, with multi-lingual staff, minimised selection bias; nurses measuring anthropometric measurements were blinded to children’s antibiotic exposure status; low proportion of missing data for antibiotic exposure and obesity (<5%); establishment of temporality as exposure was measured prior to outcome; and high-precision equipment, such as SECA scales, were used for measurement.

### Limitations

In the UK, up to 50% of prescribed medications, including antibiotics, are not taken as directed^49^, which can lead to exposure misclassification. This would lead to false positives if people do not take their prescriptions, resulting in bias toward the null. Few people (1%) report access to antibiotics without prescription^50^ making false negatives less likely

Outcome misclassification may occur due to BMI’s low sensitivity (14.3%-60%)^51^, underestimating obesity compared to body fat percentage or skinfold thickness^39,51^. However, other studies found obesity prevalence detected by body fat percentage was almost identical to BMI z-score>85^th^ percentile^39^. The latter was used to define the outcome in the sensitivity analysis. Unmeasured residual confounding could partially explain the antibiotic-obesity association. The low pseudo R^2^ suggests that a large portion of the variability in obesity remains unexplained by the model, possibly due to unmeasured factors like exercise, diet, sleep and infections (Figure 1). While UK growth charts (UK90) were used for both British Pakistani and White British children to allow for direct ethnic comparisons, it is important to acknowledge that growth trajectory differences by ethnicity have been observed^52^. Excluding multiple births limits generalisability to singleton births. Antibiotic exposure was higher in the study population, (especially in early-life (72%)) than expected UK-wide^53^. Previous BiB studies suggest high prescribing rates in Bradford are indicative of poor health rather than overprescribing^54^

### Recommendations for research

Future work should investigate the impact of specific antibiotic types, dosage and adherence to prescribed antibiotic regimens. Several studies found self-reported antibiotic use underestimates actual prescriptions or pharmacy records^55^, however, it is unclear whether this is discrepancy is due to recall bias or non-adherence. Therefore, a combination of methods is recommended to accurately capture antibiotic exposure. To address concerns about the sensitivity of BMI, future research should compare different BMI z-score thresholds with a gold standard like dual-energy X-ray absorptiometry^56^. A previous BiB study found that South Asian children had a greater mean total body fat percentage than White children within the same BMI category^57^. This reinforced the need for ethnicity-specific BMI thresholds to improve sensitivity, while maintaining high specificity in obesity classification, across different ethnicities. Finally, the association between prenatal and early-life antibiotic use and obesity in other populations, both within and outside the UK, is required to determine the generalisability of these findings.

## Conclusion

This study provides observational evidence of the association between early-life antibiotic use and increased obesity at 4-5 years in a multicultural, socioeconomically deprived UK city, consistent with previous animal and human studies. No evidence of effect modification by ethnicity (British Pakistani and White British) was found, despite the higher risk of obesity-related morbidity in South Asian ethnic origin individuals observed in previous studies^27,58^. The diverse demographics of the BiB cohort provided the statistical power to detect these associations by ethnicity, overcoming limitations of previous studies.

The findings are relevant to other UK cities with similar demographic profiles, particularly those with high levels of deprivation^59^ and South Asian populations. Further research is required to validate these findings and to explore whether this association persists into adolescence and adulthood. The results highlight the potential unintended long-term consequences of antibiotic use. This underscores the importance of cautious antibiotic prescribing practices and clear labelling of antibiotic side effects, not only to combat antibiotic resistance but also to protect against potential contributions to the growing obesity epidemic.

## Data availability

The data that support the findings of this study are available on request from the corresponding author (https://borninbradford.nhs.uk/research/how-to-access-data/). The data are not publicly available due to privacy or ethical restrictions.

## Acknowledgements

Born in Bradford is only possible because of the enthusiasm and commitment of the children and parents in BiB. We are grateful to all the participants, health professionals, schools and researchers who have made Born in Bradford happen.

