## Supplementary material for "The association between prenatal and early-life antibiotic use and obesity at 4-5 years in the Born in Bradford birth cohort"

*Supplementary material 1: Antibiotic code list*
Antibiotic code list using BNF codes that were included in the antibiotic exposure for mothers and children. See reference for more details[161].

| BNF code | Antibiotic name |
|---|---|
| 5.1.1 | Penicillins |
| 5.1.2 | Cephalosporins and other Beta-Lactams |
| 5.1.3 | Tetracyclines |
| 5.1.4 | Aminoglycosides |
| 5.1.5 | Macrolides |
| 5.1.6 | Clindamycin and Lincomycin |
| 5.1.7 | Some other antibacterial |
| 5.1.8 | Sulfonamides and Trimethoprim |
| 5.1.9 | Antituberculosis Drugs |
| 5.1.10 | Antileprotic Drugs |
| 5.1.11 | Metronidazole, Tinidazole and Ornidazole |
| 5.1.12 | Quinolones |
| 5.1.13 | Urinary-Tract Infections |

*Supplementary material 2: Methods, results, strengths and limitations for subgroup analysis with breastfeeding.*

*Methods:*

Breastfeeding was defined as any positive response to a child being breastfed to create a binary variable of any breastfeeding vs none recorded. We excluded any questions relating to feeding intention. The data was made up of a combination of questionnaire data asked in 2,048 mothers - Was child ever breastfed? Yes, No, Don’t know. For everyone who answered the questions 45% reported any breastfeeding. Routine data sources were searched for ctv3 codes relating to feeding method, and GP and Health visitor records were combined for mothers and children. We used any code at any point in the first year to define exposure. This analysis is limited to n=8991 children, 2058 (22%) had a record of any breastfeeding. We limited multivariate models to just early-life antibiotic exposure as breastfeeding is likely on the causal pathway between prenatal antibiotic use and obesity (Figure 1).

*Results:*

Cross-tabulation of breastfeeding and early-life antibiotic use (N=8,991).

| | | Ever breastfed (N=2,058) | Not breastfed (N=6,993) |
|---|---|---|---|
| Early-life antibiotic exposure: | No | 569 (22.88) | 1,918 (77.12) |
| | Yes | 1,489 (22.89) | 5,015 (77.11) |

The association between early-life antibiotic exposure and having obesity at 4-5 years (N=8,991).

| | Crude OR* (95% CI), p-value | aOR** (95% CI), p-value |
|---|---|---|
| Early-life antibiotic use (no) | 1 | 1 |
| Early-life antibiotic use (yes) | 1.378 (1.162, 1.634), <0.001 | 1.362 (1.145, 1.619), <0.001 |

Crude, aORs and 95% CIs and p-values are from logistic regression models
*Adjusted for *a priori* confounders sex and ethnicity
**Additionally adjusted for maternal pre-pregnancy obesity, birthweight, socio-economic position, smoking during pregnancy, prenatal antibiotic use and breastfeeding
Abbreviations: N, sample size; OR, odds ratio; aOR, adjusted odds ratio; CI, confidence interval.

In this population breastfeeding was associated with having obesity as children that were breastfed had 25% lower odds of having obesity at 4-5 years without adjusting for any confounders (OR 0.752; 95% CI 0.628, 0.901; p=0.002). Breastfeeding was not associated with early-life antibiotic use as the same proportion of children were ever breastfed among those that were exposed and not exposed to antibiotics in early-life. Therefore, breastfeeding does not meet the criteria to be entered as a confounder in this population. Despite this, an analysis adjusting for breastfeeding in addition to the confounders included in the main analysis confirmed the results of the main analysis with early-life antibiotic use being associated with 1.36 times the odds of having obesity at 4-5 years (95% CI 1.15, 1.62).

*Strengths:*

The addition of breastfeeding exposure to the main finding does not materially change the conclusions. The number of participants missing from the sensitivity analysis was low.

*Limitations:*

The definition of breastfeeding was crude, and we used a binary exposure. Questionnaire data was only available from a small proportion of survey data. The addition of codes from GP and health visiting data do aid the definition and improved our sample size, however information on feeding practices is not routinely collected and may be inherently biased, for example, only recorded if a health professional has concerns about the child's development. Please note the proportion reporting breastfeeding in survey data is twice that of data from routine records, and missing data is not missing at random.

*Supplementary material 3: Baseline characteristics of study population stratified by ethnicity and compared to BiB cohort.*

Distribution of baseline characteristics of all children included in BiB (N=13,685) and study population for complete case analysis (N=10,446). The latter stratified by ethnicity: Pakistani or White British

| | BiB population | | Study population | | |
|---|---|---|---|---|---|
| | Total, N = 13,685 | | Total, N = 10,446 | Pakistani, N=5,707 | WB, N=4,739 |
| Variable name | n (%) | Missing | n (%) | n (%) | n (%) |
| Child characteristics | | | | | |
| BMI: | | | | | |
| Normal/underweight | 10,851 (80.66) | | 8,359 (80.02) | 4,663 (81.71) | 3,696 (77.99) |
| Overweight | 1,396 (10.38) | | 1,117 (10.69) | 520 (9.11) | 597 (12.60) |
| Obese (at 4-5 years) | 1,206 (8.96) | 232 | 970 (9.29) | 524 (9.18) | 446 (9.41) |
| Prenatal antibiotic exposure: | | | | | |
| In the 1st trimester | 2,079 (15.41) | 190 | 1,716 (16.43) | 1,000 (17.52) | 716 (15.11) |
| In the 2nd trimester | 2,130 (15.78) | | 1,752 (16.77) | 986 (17.28) | 766 (16.16) |
| In the 3rd trimester | 1,761 (13.05) | | 1,491 (14.27) | 756 (13.25) | 735 (15.51) |
| In any trimester | 4,579 (33.93) | 190 | 3,765 (36.04) | 2,092 (36.66) | 1,673 (35.30) |
| Early-life antibiotic exposure (0-24 months) | 9,459 (70.66) | 298 | 7,563 (72.40) | 4,345 (76.13) | 3.218 (67.90) |
| Sex (male) | 7,063 (51.61) | 0 | 5,374 (51.45) | 2,927 (51.29) | 2,447 (51.64) |
| Ethnicity: | | | | | |
| Pakistani | 6,029 (45.40) | 406 | 5,707 (54.63) | — | — |
| White British | 5,059 (38.10) | | 4,739 (45.37) | — | — |
| Other | 2,191 (16.50) | | 0 | — | — |
| Preterm (<37 weeks) | 842 (6.30) | 322 | 542 (5.27) | 270 (4.78) | 282 (5.86) |
| Low birthweight (<2500g) | 1,112 (8.32) | 323 | 749 (7.28) | 488 (8.65) | 261 (5.62) |
| Child born by caesarean | 3,058 (22.94) | 355 | 2,222 (21.64) | 1,163 (20.64) | 1,059 (22.87) |
| Maternal characteristics | | | | | |

| | | | | | |
|---|---|---|---|---|---|
| Maternal smoking during pregnancy | 1,494 (10.93) | 14 | 1,200 (11.49) | 198 (3.47) | 1,002 (21.14) |
| Mother consumed alcohol in pregnancy or 3 months before | 3,480 (30.61) | 2316 | 2,870 (32.00) | 14 (0.29) | 2,856 (68.00) |
| Mother with gestational diabetes | 150 (1.12) | 233 | 114 (1.09) | 82 (1.44) | 32 (0.68) |
| Mother born outside the UK | 4,139 (36.55) | 2361 | 2,837 (31.52) | 2,765 (57.66) | 72 (1.71) |
| Mother employed at baseline | 5,005 (44.25) | 2374 | 3,813 (42.41) | 1,116 (23.31) | 2,697 (64.15) |
| Mother education higher than A-level at baseline | 2889 (27.72) | 3262 | 2,031 (24.28) | 1,234 (26.98) | 797 (21.02) |
| Primiparous at child's birth | 5,099 (39.62) | 814 | 3,797 (38.30) | 1,693 (31.23) | 2,104 (46.82) |
| Socio-economic position of household: | | | | | |
| 1 (Least deprived and most educated) | 2,231 (19.70) | 2360 | 1,537 (17.15) | 882 (18.48) | 655 (15.64) |
| 2 | 2,248 (19.85) | | 1,845 (20.59) | 448 (9.39) | 1,397 (33.35) |
| 3 | 1,721 (15.20) | | 1,336 (14.91) | 715 (14.98) | 621 (14.82) |
| 4 | 3,325 (29.36) | | 2,799 (31.23) | 2,091 (43.81) | 708 (16.90) |
| 5 (Most deprived) | 1,800 (15.89) | | 1,445 (16.12) | 637 (13.35) | 808 (19.29) |
| Mother age at child birth: | | | | | |
| 15-24 | 4,407 (32.20) | 0 | 3,481 (33.32) | 1,618 (28.35) | 1,863 (39.31) |
| 25-34 | 7,561 (55.25) | | 5,676 (54.34) | 3,396 (59.51) | 2,280 (48.11) |
| 35-49 | 1,717 (12.55) | | 1,289 (12.34) | 693 (12.14) | 596 (12.14) |
| Mother obese pre-pregnancy | 3,186 (27.87) | 2,255 | 2,641 (29.41) | 1,533 (32.05) | 1,108 (26.39) |

Abbreviations: n, number of individuals; N, sample size; BiB, Born in Bradford; BMI, body mass index.

*Supplementary material 4: Baseline characteristics of those missing antibiotic and BMI data.*

The baseline characteristics of those missing BMI and antibiotic data compared to those not missing.[a]

| Variable name | Missing BMI/antibiotic data N = 527* | Not missing BMI/antibiotic data N=12,803* |
|---|---|---|
| Child characteristics | | |
| Sex (male) | 281 (53.32) | 6,607 (51.61) |
| Ethnicity: | | |
| Pakistani | 151 (31.86) | 5,707 (45.78) |
| White British | 188 (39.66) | 4,739 (38.02) |
| Other | 135 (28.48) | 2,019 (16.20) |
| Preterm (<37 weeks) | 37 (8.04) | 672 (5.34) |
| Low birthweight (<2500g) | 30 (6.52) | 914 (7.26) |
| Child born by caesarean | 115 (25.00) | 2,775 (22.10) |
| Maternal characteristics | | |
| Mother smoked during pregnancy | 44 (8.56) | 1,420 (11.09) |
| Mother consumed alcohol in pregnancy or 3 months before: | | |
| Yes | 143 (34.96) | 3,252 (30.77) |
| No | 266 (65.04) | 7,309 (69.16) |
| Don’t remember | 0 (0) | 7 (0.07) |
| Mother with gestational diabetes | 2 (0.60) | 140 (1.10) |
| Mother born outside the UK | 160 (38.65) | 3,870 (36.50) |
| Mother employed at baseline | 177 (42.86) | 4,688 (44.27) |
| Mothers education higher than A-level | 144 (37.50) | 2,676 (27.41) |
| Primiparous at child's birth | 190 (42.60) | 4,777 (39.40) |
| Mother receives child benefits | 174 (42.34) | 5,669 (53.64) |
| Socio-economic position of household: | | |

| | | |
|---|---|---|
| 1 (Least deprived and most educated) | 120 (29.56) | 2,043 (19.40) |
| 2 | 59 (14.53) | 2,125 (20.18) |
| 3 | 52 (12.81) | 1,611 (15.30) |
| 4 | 104 (25.62) | 3,082 (29.27) |
| 5 (Most deprived) | 71 (17.49) | 1,668 (15.84) |
| Mother age at child birth: | | |
| 15-24 | 117 (33.59) | 4,137 (32.31) |
| 25-34 | 288 (54.65) | 7,071 (55.23) |
| 35-49 | 62 (11.76) | 1,595 (12.46) |
| Mother obese pre-pregnancy | 85 (20.88) | 2,960 (28.00) |

*Out of all alive singleton births in the BiB population (N=13,330)
Abbreviations: BMI, body mass index; N, sample size.

[a]Those missing BMI and/or antibiotic data were more likely to be preterm; their mothers were more likely to have higher education, less likely to have smoked during pregnancy, and less likely to be born in the UK (supplementary material 3). The proportion of missing antibiotic data was similar across obese and non-obese children and among other explanatory variables. The proportion of children missing BMI data was also similar across exposed and unexposed groups and other explanatory variables. Those missing ethnicity data generally lacked data on all baseline covariates. For the remaining explanatory variables and exposure/outcome, the proportion of missing ethnicity data was similar across all groups. The baseline questionnaire had an 83% response rate, resulting in approximately 20% missing data for all the variables it measured. The proportion of missing data was similar across exposure and outcome groups. Missing questionnaire data was more common in women with diabetes, multiparous women, Pakistani, and mothers of preterm, low birthweight or caesarean children.

*Supplementary material 5: Sensitivity analysis.*

The association between any prenatal (Model 1) and any early-life (Model 3) antibiotic use and obesity at 4-5 years with stratification by ethnicity and sex, among 1st siblings (N=9,371)

| | Sensitivity analysis 1: Excluding 2nd and 3rd siblings | | | | | |
|---|---|---|---|---|---|---|
| | Model 1 prenatal | | | Model 3 early-life | | |
| | Crude OR* (95% CI), p-value | aOR** (95% CI), p-value | interaction p-value[i] | Crude OR* (95% CI), p-value | aOR*** (95% CI), p-value | interaction p-value[i] |
| All | 1.174 (1.019, 1.354), 0.027 | 1.096 (0.940, 1.279), 0.243 | – | 1.419 (1.200, 1.678), <0.0001 | 1.438 (1.1198, 1.725), 0.0001 | – |
| Stratified by ethnicity | | | | | | |
| White British | 1.238 (1.006, 1.523), 0.045 | 1.093 (0.874, 1.368), 0.437 | 0.8885 | 1.386 (1.101, 1.743), 0.005 | 1.420 (1.109,1.817), 0.005 | 0.8899 |
| Pakistani | 1.122 (0.923, 1.363), 0.250 | 1.088 (0.879, 1.346), 0.440 | | 1.457 (1.141, 1.862), 0.002 | 1.474 (1.123, 1.934), 0.004 | |
| Stratified by sex | | | | | | |
| Female | 1.101 (0.896, 1.351), 0.362 | 0.965 (0.771, 1.207), 0.755 | 0.2036 | 1.521 (1.202, 1.923), 0.0003 | 1.655 (1.276, 2.146), 0.0001 | 0.1847 |
| Male | 1.248 (1.025, 1.519), 0.029 | 1.226 (0.991, 1.517), 0.062 | | 1.318 (1.039, 1.673), 0.020 | 1.260 (0.975, 1.627), 0.072 | |

Crude and aORs with 95% CIs are from logistic regression models; P-values are from Likelihood Ratio Tests testing the null hypothesis of no association.

*Crude models are adjusted for *a priori* confounders.
**Adjusted for *a priori* cofounders, diabetes, maternal pre-pregnancy obesity, socio-economic position, smoking during pregnancy.
***Adjusted for *a priori* confounders, maternal pre-pregnancy obesity, birthweight, socio-economic position, smoking during pregnancy, antibiotic use during pregnancy.

[i]p-value from formal Likelihood Ratio Tests for interaction, testing effect modification

Abbreviations: N, sample size; OR, odds ratio; aOR, adjusted odds ratio; CI, confidence interval; BMI, body mass index

The association between any prenatal (Model 1) and any early-life (Model 3) antibiotic use and overweight/obesity at 4-5 years with stratification by ethnicity and sex (N=10,446)

| Sensitivity analysis 2: Outcome is defined as overweight or obese (BMI z-score>85th percentile) | | | | | | |
|---|---|---|---|---|---|---|
| | Model 1 prenatal | | | Model 3 early-life | | |
| | Crude OR* (95% CI), p-value | aOR** (95% CI), p-value | interaction p-value[i] | Crude OR* (95% CI), p-value | aOR*** (95% CI), p-value | interaction p-value[i] |
| All | 1.137 (1.031, 1.256), 0.011 | 1.111 (0.997, 1.238), 0.056 | — | 1.215 (1.087, 1.357), 0.0005 | 1.215 (1.076, 1.372), 0.002 | — |
| Stratified by ethnicity | | | | | | |
| White British | 1.048 (0.908, 1.210), 0.519 | 0.969 (0.830, 1.132), 0.691 | 0.02 | 1.173 (1.009, 1.363), 0.036 | 1.224 (1.040,1.440), 0.014 | 0.876 |
| Pakistani | 1.228 (1.070, 1.409), 0.004 | 1.257 (1.081, 1.463), 0.003 | | 1.267 (1.074, 1.494), 0.004 | 1.215 (1.011, 1.460), 0.036 | |
| Stratified by sex | | | | | | |
| Female | 1.075 (0.933, 1.241), 0.317 | 1.040 (0.890, 1.215), 0.624 | 0.273 | 1.207 (1.035, 1.407), 0.015 | 1.232 (1.041, 1.460), 0.015 | 0.912 |
| Male | 1.199 (1.045, 1.376), 0.010 | 1.183 (1.018, 1.374), 0.029 | | 1.224 (1.042, 1.437), 0.013 | 1.200 (1.008, 1.430), 0.039 | |

Crude and aORs with 95% CIs are from logistic regression models; P-values are from Likelihood Ratio Tests testing the null hypothesis of no association.

*Crude models are adjusted for a *priori* confounders.

**Adjusted for *a priori* cofounders, diabetes, maternal pre-pregnancy obesity, socio-economic position, smoking during pregnancy.
***Adjusted for *a priori* confounders, maternal pre-pregnancy obesity, birthweight, socio-economic position, smoking during pregnancy, antibiotic use during pregnancy.

[i]p-value from formal Likelihood Ratio Tests for interaction, testing effect modification

Abbreviations: N, sample size; OR, odds ratio; aOR, adjusted odds ratio; CI, confidence interval; BMI, body mass index
